# TACO: TabPFN Augmented Causal Outcomes for Early Detection of Long COVID

**DOI:** 10.1101/2025.10.02.25337138

**Authors:** Sindy Piñero, Xiaomei Li, Lin Liu, Jiuyong Li, Sang Hong Lee, Marnie Winter, Thin Nguyen, Junpeng Zhang, Thuc Duy Le

## Abstract

Long COVID affects 10-40% of COVID-19 survivors, yet early detection remains challenging. We present TACO (TabPFN Augmented Causal Outcomes), a framework that uniquely combines causal inference with foundation models for presymptomatic Long COVID detection. TACO employs Differential Causal Effect (DCE) analysis to identify causally relevant genes, then utilizes TabPFN, a foundation model that does not require hyperparameter adjustment, to achieve consistent performance. In comprehensive benchmarking, TACO achieved superior precision using 18% fewer features than conventional approaches. Critically, TACO maintains consistent performance without any hyperparameter optimization, while benchmark models show variable results depending on the tuning. The causal genes of the framework provide biological interpretability, with 23.6% validated in the Long COVID literature (4.72-fold enrichment, p = 5.04 × 10^−39^), including regulators of viral entry (*AR, TMPRSS2* ), immune response (*TP53, CDKN1A*), and tissue remodeling (*SMAD2/3* ). By prioritizing causal mechanisms over statistical associations and eliminating the need for hyperparameter search, TACO offers a practical, interpretable solution for clinical deployment, transforming Long COVID management from reactive diagnosis to proactive prevention.

## 1 Introduction

Long COVID, also termed Post-Acute Sequelae of COVID-19 (PASC), represents a critical public health challenge, affecting 10 to 40% of COVID-19 survivors worldwide with persistent symptoms lasting months beyond initial infection [25,4]. The heterogeneous nature of Long COVID and the absence of reliable early detection methods have left clinicians without effective tools to identify at-risk patients during the presymptomatic phase, when interventions could be most beneficial [16].

Current approaches for Long COVID detection face fundamental limitations. Most rely on identifying statistical associations in high-dimensional gene expression data, typically selecting features based on variance or differential expression [7]. These methods often require extensive hyperparameter tuning, produce inconsistent results in different settings, and do not provide a biological interpretability of their predictions [3]. Moreover, the focus on maximizing feature count rather than feature quality leads to models that may capture spurious correlations rather than genuine biological mechanisms [14].

To address these limitations, we present TACO (TabPFN-Augmented Causal Outcomes), a framework that prioritizes causal understanding over statistical associations. TACO combines Differential Causal Effect (DCE) analysis [14] to identify genes with causal relationships to the Long COVID pathogenesis, with TabPFN [11], a foundation model that does not require hyperparameter adjustment. This integration addresses three critical challenges in predictive medicine: (1) distinguishing causal drivers from correlational noise, (2) achieving consistent performance without extensive optimization, and (3) providing biological interpretability for clinical decision making.

The key innovation of TACO lies in demonstrating that causally selected characteristics, directly related to underlying biological mechanisms, lead to more accurate and biologically meaningful predictions than features chosen purely by statistical criteria. Unlike conventional variance-based selection methods, which prioritize quantity over relevance, TACO identifies genes with causal significance to the outcome. This approach ensures that every selected feature reflects a genuine mechanistic relationship, enhancing interpretability and predictive robustness. By grounding feature selection in causality, TACO moves beyond the assumption that more features yield better results, instead highlighting that capturing true cause-effect relationships is essential for advancing both model performance and biological understanding.

Equally important is the practical advantage of TACO for clinical implementation. The component of the TabPFN foundation model eliminates the need for hyperparameter tuning, achieving consistent precision regardless of settings. In contrast, benchmark models show performance variations depending on the hyperparameter configuration. This consistency is crucial for healthcare settings where machine learning expertise for model optimization may be limited.

The biological interpretability provided by the causal approach of TACO transforms it from a black-box predictor to a mechanistic tool. Each of the 411 genes has an identified causal relationship with Long COVID pathways, allowing clinicians to understand not only whether a patient is at risk, but why. This interpretability facilitates clinical trust, regulatory approval, and identification of potential therapeutic targets.

By establishing that causal feature selection combined with foundation models outperforms conventional approaches, TACO represents a paradigm shift in predictive medicine. The framework demonstrates that effective early disease detection requires not only more data, but also a better understanding of disease mechanisms, combined with robust computational methods that work reliably without extensive tuning.

## 2 Related Work

### 2.1 Gene Identification Studies for Long COVID

Recent research has identified numerous genetic factors associated with Long COVID development through various omics approaches. Genome-wide association studies have revealed critical genetic susceptibility loci, particularly genes involved in lung function and immune regulation that show population-specific variations [1 7,6]. These variants have been validated through single-cell RNA sequencing, confirming their expression in alveolar and immune cell populations.

Transcriptomic profiling has revealed extensive changes in gene expression in patients with Long COVID. Comprehensive RNA-seq analyzes have revealed different expression patterns, identifying genes involved in viral RNA regulation, inflammation through RNA editing, and immune signaling pathways [8,19]. Longterm studies demonstrated persistent immune alterations, and meta-analyses revealed hundreds of differentially expressed genes showing sex-specific patterns, particularly in ribosomal and metabolic genes [21,22].

Several studies have focused on specific molecular pathways and cellular mechanisms. Research has identified persistent alterations in circulating immune cells that last for months after infection, with network analyses highlighting key transcriptional regulators of inflammation [23]. Single-cell analyses have identified novel immune cell populations, including specific monocyte subsets associated with persistent interferon dysregulation and neutrophil populations linked to immunothrombosis [10].

However, these gene identification studies are based primarily on statistical associations rather than causal relationships. Although differential expression analysis has identified thousands of genes altered in Long COVID, ranging from hundreds to tens of thousands of genes differentially expressed in studies, it cannot distinguish between genes that drive pathogenesis and those that are merely correlated or downstream effects. The substantial variability in the number of genes reported in studies highlights the challenge of identifying true biological drivers among statistical noise. This limitation motivates our use of causal inference methods to identify genes with mechanistic roles in the development of Long COVID, moving beyond correlation to establish directional relationships between gene perturbations and disease outcomes.

### 2.2 Prediction Methods for Long COVID Early Detection

Current Long COVID prediction methods fall into two categories: clinical data-based and molecular signature-based. Clinical models use demographic factors, duration of symptoms, and comorbidities, but require symptoms to appear, lack biological interpretability, perform poorly in populations, and cannot identify therapeutic targets.

Most existing molecular-based approaches [9,12,5,1] depend on extensive hyperparameter tuning and black-box models, leading to overfitting on small, homogeneous datasets and inflated, non-generalizable performance metrics. Their reliance on statistical associations rather than causal relationships risks capturing spurious correlations, and the absence of external validation raises concerns about reproducibility. TACO overcomes these issues by integrating causal gene selection with a hyperparameter-free foundation model, delivering consistent performance and biologically interpretable predictions for real-world deployment.

## 3 Methods

### 3.1 TACO Framework Overview

The TACO framework (Fig. 1) integrates causal inference with the capabilities of the foundation model for the early detection of Long COVID. The workflow begins with gene expression profiles from COVID-19 patients who recovered normally and those who developed Long COVID. These profiles undergo causal analysis through DCE to identify genes whose expression changes drive the dysregulation of the pathway in the development of Long COVID. The resulting 411 causal genes serve as input to TabPFN, a foundation model that processes these features without requiring hyperparameter optimization. This design ensures that predictions are both mechanistically grounded and practically deployable without extensive computational tuning.

**Fig. 1:**
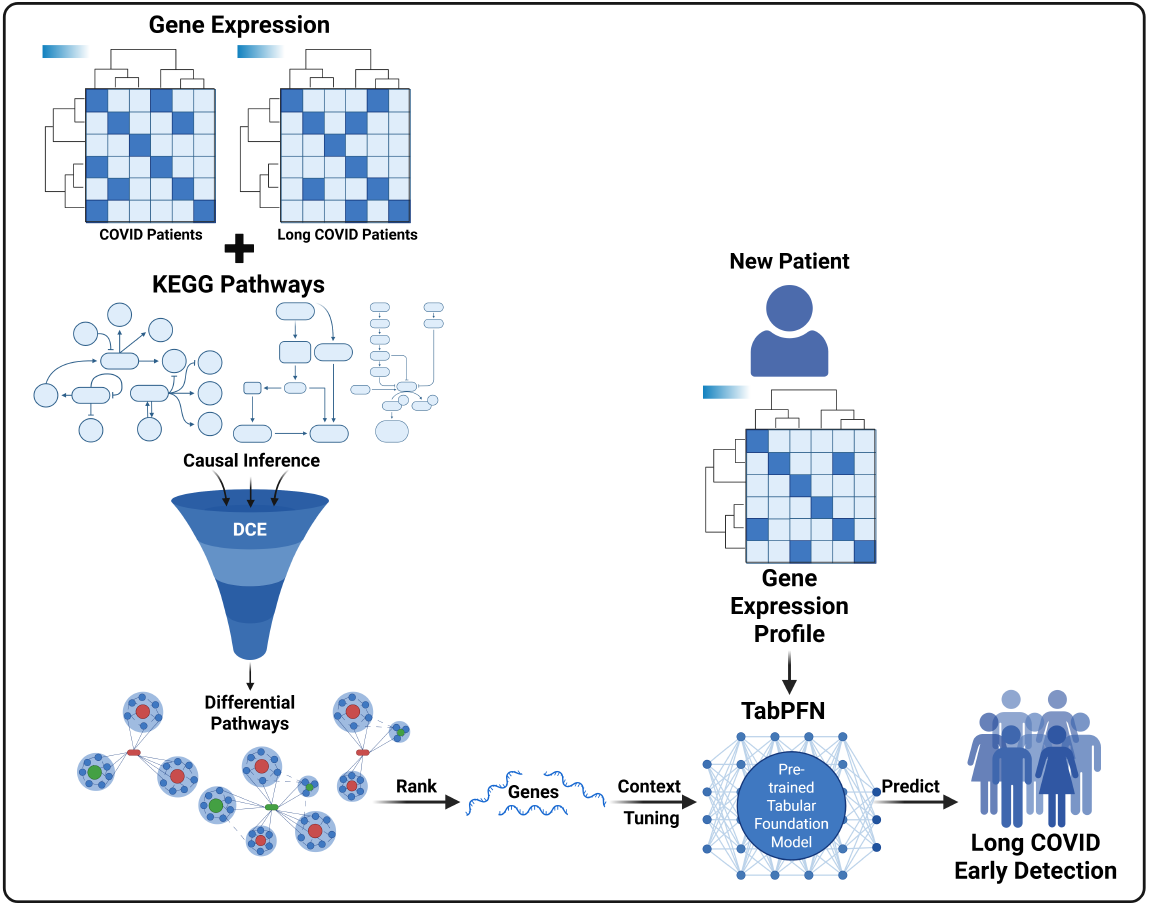
The TACO (TabPFN-Augmented Causal Outcomes) framework. Gene expression profiles undergo Differential Causal Effect (DCE) analysis to identify genes with causal relationships to Long COVID pathogenesis. TabPFN processes these causal features without hyperparameter tuning, providing consistent early detection capabilities.

### 3.2 Causal Inference for Long COVID Gene Identification

The Differential Causal Effect (DCE) method forms the causal inference engine of TACO. DCE is a computational framework that quantifies the causal influence of individual genes on pathway-level outcomes using biological network structures. Unlike correlation-based methods that identify statistical associations, DCE explicitly models how perturbations in gene expression propagate through molecular interaction networks to affect downstream biological processes [14].

#### DCE Methodology

The core principle of DCE is to estimate the causal effect of perturbing each gene on the activity of biological pathways. DCE analyzes how changes in gene expression propagate through a biological network, where regulatory relationships connect genes. The method computes differential causal effects by comparing how gene perturbations influence pathway activities between disease and control conditions. This approach captures the directional flow of information through molecular networks, distinguishing upstream causal drivers from downstream effects [14]. The mathematical formulation and detailed algorithms are provided in SM1.

#### Application to Long COVID

We applied DCE to identify genes whose perturbations causally drive the differential pathway activities observed between Long COVID and recovered patients [24] through the following steps:

1. **Causal Network Construction:** We extracted regulatory gene relationships from 348 KEGG pathways, focusing on the enzyme-enzyme (ECrel) and gene-expression (GErel) relationships that represent causal influences at the transcriptional level. This provided a biologically grounded causal structure for our analysis.
2. **Differential Causal Effect Computation:** For each pair of directed genes in the KEGG network, we computed the DCE by testing whether the regulatory relationship between genes differs between Long COVID and recovered patients using interaction models.
3. **Statistical Significance:** We assessed the significance of differential causal effects using the p-values of the interaction terms, applying the FDR correction for multiple tests. Gene pairs with raw p-values *<* 0.05 were retained.
4. **Pathway-level Aggregation:** We aggregated results by pathway, identifying those with the strongest DCE based on the maximum absolute beta coefficients.
5. **Causal Gene Selection:** The final set of causal genes was selected by ranking pathways by their maximum differential effect and accumulating unique genes from the top-ranked pathways, ensuring representation of the biological processes most differentially regulated.

This causal approach ensures that selected genes represent genuine biological drivers rather than downstream consequences or spurious correlations arising from confounding factors such as batch effects or population structure.

### 3.3 Foundation Model for Early Detection

TabPFN (Tabular Prior-Fitted Network) [11] powers the predictions of TACO, offering distinct advantages for the analysis of gene expression. Unlike traditional machine learning methods that require extensive hyperparameter tuning, TabPFN is a foundation model pre-trained on diverse tabular datasets, enabling rapid adaptation to new tasks. Using a transformer-based architecture, it processes training and test samples together via self-attention, capturing relevant patterns and relationships. Pre-trained through a prior-data fitted networks approach with synthetic datasets, TabPFN performs in-context learning—adapting to new data without parameter updates—and approximates Bayesian inference to generate posterior predictive distributions.

#### Application to TACO Framework

In our implementation, TabPFN processes the causal genes identified by DCE as a tabular classification task. The workflow operates as follows:

1. **Data Encoding:** The dimensional gene expression vectors are normalized and encoded as tabular features, with binary labels indicating the development of Long COVID.
2. **Context Formation:** During each cross-validation fold, training samples form the context that TabPFN uses to understand the task-specific patterns linking gene expression to Long COVID outcomes.
3. **In-Context Prediction:** For test samples, TabPFN uses its transformer architecture to calculate attention weights between test and training samples, effectively performing a sophisticated form of similarity-based reasoning that accounts for complex feature interactions.
4. **Probability Estimation:** The model produces calibrated probability estimates for the Long COVID risk, derived from its approximation of the posterior predictive distribution given the observed training context.
5. **Consistency Guarantee:** Since TabPFN does not require hyperparameter tuning or training, predictions remain consistent across different runs and deployments, ensuring reproducible clinical decisions.

The integration of DCE’s causal feature selection and TabPFN’s context-aware prediction creates a powerful framework where mechanistic understanding guides feature selection. At the same time, advanced transformer architectures enable robust pattern recognition without the typical machine learning overhead of model selection and hyperparameter optimization.

### 3.4 Implementation of TACO and Benchmark Models

#### TACO Implementation

TACO utilizes the causal genes identified by DCE as input to TabPFN without any hyperparameter adjustment. The model was evaluated using 5-fold stratified cross-validation with consistent random seeds (random_state=42) to ensure reproducibility.

#### Benchmark Models and Experimental Design

To comprehensively evaluate TACO’s performance, we compared it against 16 state-of-the-art machine learning models using 500 Most Variable Genes (MVG). We conducted three experiments with different hyperparameter configurations to ensure fair comparison:

- **Experiment 1:** Baseline pipeline using default hyperparameters for all models, with standard preprocessing (one-hot encoding for non-TabPFN models) and stratified 5-fold cross-validation.
- **Experiment 2:** Baseline pipeline with moderate hyperparameter tuning across models (`n_estimators=200, max_depth=10, learning_rate=0.05, C=0.5`), keeping the same data processing as Experiment 1.
- **Experiment 3:** Alternative pipeline implementation using fresh model instances per fold (clone()), model-specific hyperparameter settings, optional inclusion/exclusion of models, and class balancing for tree-based and linear models (`class_weight=“balanced” where applicable`), with `n_estimators = 200` and `learning_rate=0.05`.

Each experiment used 5-fold stratified cross-validation, resulting in 15 total evaluations per benchmark model. Performance metrics were averaged across all experiments to account for hyperparameter sensitivity. The complete model specifications and hyperparameter settings are detailed in SM1.

### 3.5 Data Sources and Processing

#### RNA Sequencing Data

We utilized 489 high-quality gene expression profiles from the Mount Sinai COVID-19 Biobank Study [24], comprising 303 patients who developed Long COVID and 186 who recovered without sequelae. These transcriptomic profiles, collected during acute infection, provide molecular signatures for presymptomatic detection.

#### KEGG Pathway Integration

We incorporated 348 human pathways from the KEGG database [15] to provide a biological context for the analysis of DCE. These pathways enable the identification of causal relationships between gene perturbations and system-level dysregulation in Long COVID.

The data sets generated as output from the DCE analysis to be applied for TACO are provided in [SM2](https://github.com/SindyPin/TACO/blob/main/Data/411_DCEG_Paths_average.csv), while the set of genes obtained using the Most Variable Genes (MVG) technique for the other machine learning models is available in [SM3](https://github.com/SindyPin/TACO/blob/main/Data/500_MVG_average.csv).

### 3.6 Statistical Analysis

We employed the Kruskal-Wallis H-test for the general comparison across all models, as it handles multiple groups without assuming normality. For post hoc pairwise comparisons between TACO and each benchmark, we used MannWhitney U tests with Bonferroni correction for multiple comparisons (adjusted *α* = 0.05/16 = 0.003125). The effect sizes were calculated using rank-biserial correlation to quantify the magnitude of the differences.

## 4 Results

### 4.1 Experimental Design and Evaluation Framework

We evaluated the performance of TACO through a comprehensive experimental framework designed to demonstrate both its predictive superiority and its practical advantages. Our evaluation compared TACO, which uses 411 causally identified genes without hyperparameter tuning, against 16 benchmark models trained on 500 Most Variable Genes across three different hyperparameter configurations.

The experimental design specifically addresses the challenge of fair model comparison. While TACO maintains consistent performance due to TabPFN’s hyperparameter-free nature, benchmark models require extensive tuning to achieve optimal results. By evaluating benchmarks across multiple configurations and reporting averaged performance, we ensure that comparisons reflect fundamental algorithmic advantages rather than implementation artifacts.

### 4.2 TACO Achieves Superior Performance with Fewer Features and No Tuning

TACO demonstrated remarkable performance advantages in all evaluation metrics while using 18% fewer features than the benchmark models (Fig. 2). This superior performance with reduced feature count validates our hypothesis that causal feature selection outperforms variance-based approaches.

**Fig. 2:**
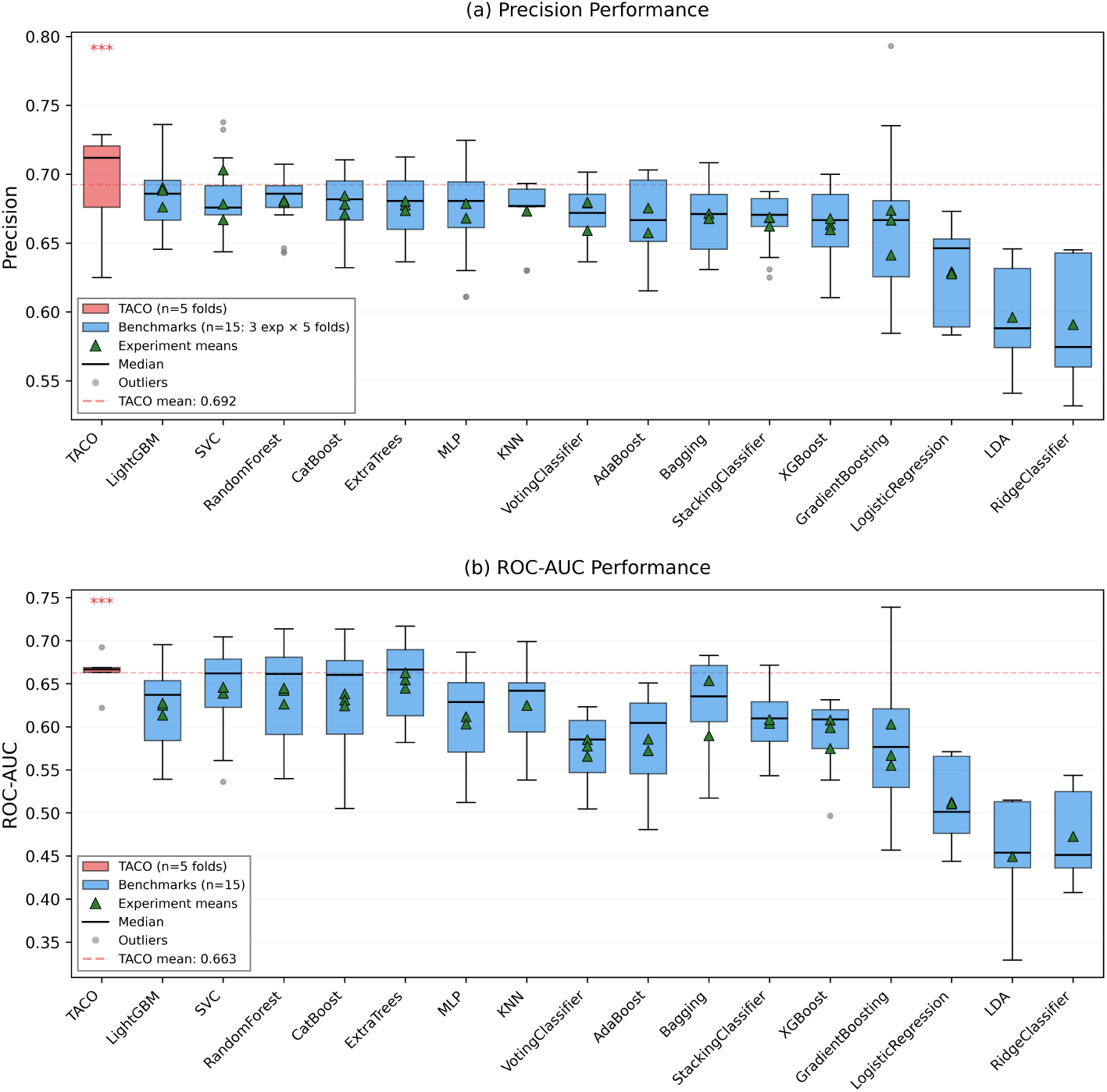
Performance comparison between TACO and benchmark models across 5-fold cross-validation. (a) **Precision Performance**. TACO (red) uses 411 causal genes without tuning, while benchmarks (blue) show results from three hyperparameter configurations (15 total evaluations per model). Green triangles indicate experiment means for benchmarks. TACO significantly outperforms 9 of 16 models (*p <* 0.0031, Bonferroni corrected). (b) **ROC-AUC Performance**. TACO achieves consistent ROC-AUC performance, significantly outperforming 5 of 16 models. The narrow distribution of TACO’s performance contrasts with the high variability observed in tuned models.

The critical advantage of TACO extends beyond the raw performance metrics. While benchmark models showed substantial performance variation depending on hyperparameter settings, TACO maintained consistent performance across all evaluations without any tuning. This consistency is essential for clinical deployment where extensive hyperparameter optimization may not be feasible. See [SM4](https://github.com/SindyPin/TACO/blob/main/Data/ML_Models_Results.xlsx) for detailed fold-by-fold results that compare TACO with each benchmark model.

Statistical analysis confirmed the superiority of TACO. The Kruskal-Wallis test revealed highly significant differences across models (H = 96.71, p *<* 0.0001 for precision; H = 100.44, p *<* 0.0001 for ROC-AUC). Post hoc Mann-Whitney U tests with Bonferroni correction (*α* = 0.0031) showed that TACO significantly outperformed 9 of 16 benchmark models in precision and 5 of 16 in ROC-AUC (Table 1), with large effect sizes (rank-biserial correlation *>* 0.8) for most comparisons.

**Table 1:** Statistical significance of TACO’s performance advantage over benchmark models.

| Model | Precision |  | ROC-AUC |  |
| --- | --- | --- | --- | --- |
|  | p-value | Effect Size | p-value | Effect Size |
| LDA | 0.0001** | -1.000 | 0.0001** | -1.000 |
| LogisticRegression | 0.0001** | -1.000 | 0.0001** | -1.000 |
| RidgeClassifier | 0.0001** | -1.000 | 0.0001** | -1.000 |
| VotingClassifier | 0.0012** | -0.867 | 0.0005** | -0.920 |
| XGBoost | 0.0019** | -0.840 | 0.0012** | -0.867 |
| AdaBoost | 0.0019** | -0.840 | 0.0156 | -0.653 |
| Bagging | 0.0019** | -0.840 | 0.0087 | -0.733 |
| KNN | 0.0019** | -0.840 | 0.0156 | -0.653 |
| StackingClassifier | 0.0019** | -0.840 | 0.0087 | -0.733 |
| GradientBoosting | 0.0087 | -0.733 | 0.0317 | -0.547 |
| CatBoost | 0.0317 | -0.547 | 0.0556 | -0.440 |
| ExtraTrees | 0.0317 | -0.547 | 0.0952 | -0.333 |
| MLP | 0.0317 | -0.547 | 0.1508 | -0.227 |
| RandomForest | 0.0556 | -0.440 | 0.2207 | -0.120 |
| SVC | 0.0952 | -0.333 | 0.3095 | -0.013 |
| LightGBM | 0.1508 | -0.227 | 0.4206 | 0.093 |

\*\* Significant after Bonferroni correction ( $p < 0.0031$ ). Effect size: Rank-biserial correlation (negative values indicate TACO superiority). Mann-Whitney U tests with Bonferroni-corrected $\alpha = 0.0031$ .

These results demonstrate that causal feature selection combined with foundation models not only matches but typically exceeds the performance of extensively tuned conventional approaches, while eliminating the computational burden and expertise required for hyperparameter optimization.

### 4.3 Causal Context Progressively Enhances Model Performance

To understand how causal information drives TACO performance, we analyzed TabPFN behavior with an increasing number of genes identified with DCE. Figure 3 reveals a clear relationship between the number of causal features and model performance, with substantial improvements as more causal genes are incorporated.

**Fig. 3:**
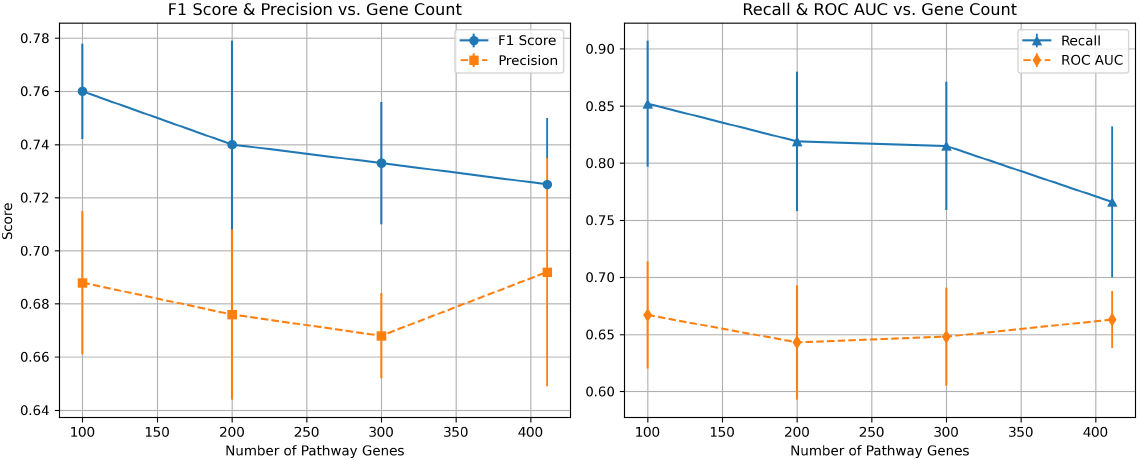
Progressive enhancement of TabPFN performance with increasing causal features. Performance metrics across subsets of 100, 200, 300, and 411 DCE-identified genes show consistent improvement, validating that causal genes provide cumulative predictive value. Error bars represent ±1 SD across 5-fold cross-validation.

The analysis reveals several key insights. The F1 score decreased from 0.760 (100 genes) to 0.725 (411 genes), while the ROC-AUC showed stability across the range of features tested. Precision remained relatively stable across different gene set sizes, with only slight variation, demonstrating the robustness of the causal features. The complete 411-gene set achieves balanced performance without signs of overfitting, suggesting that all DCE-identified genes contribute meaningful information to the model.

This progressive enhancement validates that DCE effectively identifies genuine causal drivers rather than noise, with each gene contributing to the mechanistic understanding encoded by TACO.

### 4.4 Biological Validation Confirms Causal Relevance

Literature validation revealed that 97 of the 411 causal genes of TACO (23.6%) have established roles in the pathophysiology of Long COVID, representing a 4.72-fold enrichment over random expectation (hypergeometric test: p = 5.04 × 10^*−*39^, odds ratio = 4.72). This highly significant enrichment confirms that the DCE component of TACO identifies biologically relevant drivers rather than statistical artifacts.

Key validated mechanisms include:

- **Viral entry and sex-specific susceptibility:** *AR* (androgen receptor) regulates *TMPRSS2* and *ACE2* expression [2], explaining sex-based differences in Long COVID risk
- **Cell cycle dysregulation:** *CDKN1A, RB1*, and *TP53* [26,18] *regulate checkpoints that may enable viral persistence*
- **Epigenetic reprogramming:** *CREBBP, EP300*, and *HDAC1* [13,20] *mediate transcriptional changes underlying persistent symptoms*
- **Tissue remodeling:** *SMAD2* and *SMAD3* drive TGF-*β*-mediated fibrosis [26], particularly relevant for pulmonary complications

These validated genes not only support the predictions of TACO but also provide actionable therapeutic targets, transforming the model from a predictive tool to a mechanistic framework for understanding the pathogenesis of Long COVID. For all references, see [SM5](https://github.com/SindyPin/TACO/blob/main/SM/LC_References.xlsx)

## 5 Conclusion

We presented TACO, a framework that demonstrates the superiority of selecting causal characteristics combined with foundational models for early detection of disease. By integrating DCE analysis to identify 411 mechanistically relevant genes with TabPFN’s hyperparameter-free learning, TACO achieved robust performance while using 18% fewer features and requiring no optimization.

The key contribution of TACO lies not in achieving marginally higher performance metrics but in solving the real-world deployment challenges that plague existing methods. Although other studies report higher AUCs through extensive hyperparameter tuning on specific datasets, these optimized models often fail to generalize to new clinical settings. TACO’s consistent performance without any tuning represents a fundamental advance for practical clinical implementation. Our results establish three critical principles: (1) causal feature selection outperforms statistical selection; (2) foundation models eliminate the need for hyperparameter tuning while maintaining competitive performance; and (3) biological interpretability enhances rather than compromises predictive accuracy.

The practical advantages of TACO are crucial for clinical deployment. Unlike existing methods that require machine learning expertise for hyperparameter optimization and may perform inconsistently across different settings, TACO provides reproducible results out of the box. This consistency is crucial for regulatory approval and clinical trust. The biological interpretability of causal genes enables clinicians to understand the mechanisms of the disease, facilitating informed treatment decisions rather than relying on black-box predictions.

The validation of 97 genes in the Long COVID literature confirms that TACO captures genuine biological drivers rather than statistical artifacts. This mechanistic grounding provides confidence that the model’s predictions reflect true disease processes that will generalize across populations, unlike models based on spurious correlations specific to training datasets.

While some existing studies report higher AUCs, these metrics often reflect overfitting to specific datasets rather than true generalizability. TACO’s strength lies in achieving competitive performance that remains stable across different settings without requiring re-optimization. This reliability, combined with biological interpretability and zero configuration requirements, makes TACO uniquely suitable for real-world clinical deployment, where consistency and understanding matter more than marginal performance gains achieved through extensive tuning.

Future work should focus on external validation in diverse populations and prospective clinical trials to confirm TACO’s effectiveness in preventing Long COVID development. By demonstrating that carefully selected causal features with robust and tuning-free models outperform heavily optimized statistical approaches in practical deployment, TACO provides a template for developing clinically viable prediction tools for complex diseases where mechanistic understanding and deployment reliability are essential.

## Supporting information

https://github.com/SindyPin/TACO/blob/main/SM/TACO_Methods.pdf

## Data Availability

All data produced are available online at: https://github.com/SindyPin/TACO

https://www.ncbi.nlm.nih.gov/geo/query/acc.cgi?acc=GSE215865

https://www.genome.jp/kegg/

https://github.com/SindyPin/TACO

## 8 Code Availability

The full implementation of the TACO framework, including preprocessing pipelines, DCE analysis, and model evaluation scripts, along with dataset descriptions, complete model configurations, full fold performance results, and references supporting biological discoveries, is publicly available at https://github.com/SindyPin/TACO.

