## Supplementary material for "TACO: TabPFN Augmented Causal Outcomes for Early Detection of Long COVID": https://github.com/SindyPin/TACO/blob/main/SM/TACO_Methods.pdf

#### 1 Data Preprocessing Pipeline

##### 1.1 RNA-seq Data

From the raw RNA-seq dataset, we:

1. Removed rows and columns containing only NA values.
2. Transposed the table so genes became columns and samples became rows.
3. Extracted `Subject_ID` from sample names (splitting at the first "T").
4. Averaged gene expression values across multiple samples per subject.

This process ensured one representative expression profile per subject while preserving the underlying biological variation.

##### 1.2 Clinical Data Processing

We merged current and historical symptom variables (e.g., `Symptom_X` and `Symptom_X_Ever`) so that a symptom was marked positive if present in either column. Long COVID status was assigned based on:

– Presence of at least one persistent post-COVID symptom.
– Recovery status marked as “Not Recovered”.
– Post-COVID health reported as “Worse”.

We then merged the clinical labels with the RNA-seq dataset using `Subject_ID` as the key.

##### 1.3 Most Variable Genes (MVG)

For benchmark models, we selected the 500 genes with the highest variance across subjects:

$$\text{Variance}(g_i) = \frac{1}{n-1}\sum_{j=1}^{n}(x_{ij} - \bar{x}_i)^2$$

This standard feature selection approach maximizes statistical variation but does not incorporate causal information.

#### 2 Differential Causal Effects (DCE) Implementation for TACO

The DCE framework [2] identifies genes whose expression changes causally influence Long COVID development, forming the foundation of TACO's predictive capability.

##### 2.1 DCE Analysis Pipeline

1. **Pathway Network Construction:** Extracted gene-gene interactions from KEGG [3] using:
   - ECrel (enzyme-enzyme relationships): Direct enzymatic interactions
   - GErel (gene expression relationships): Regulatory relationships
   - Excluded PPrel (protein-protein interactions) as they occur post-transcriptionally
2. **Causal Effect Estimation:** Using RNA-seq data from 1,392 samples (COVID-19 and Long COVID patients), we:
   (a) Estimated causal coefficients ($\beta$) for each source-target gene pair within pathways.
   (b) Quantified how regulatory relationships differ between disease states.
   (c) Applied FDR correction (q $<$ 0.05) for multiple testing.
3. **Gene Prioritization for TACO:** The DCE analysis identified 411 genes with significant causal effects on Long COVID pathogenesis:

$$G_{TACO} = \{g_i : |\beta_i| > \tau, \text{FDR}_i < 0.05\}$$

   where $\tau$ is the effect size threshold determined by biological significance.

These 411 genes represent:

- Genes with the strongest causal effects on pathway dysregulation.
- Molecular drivers distinguishing Long COVID from normal recovery.
- Biologically interpretable targets for intervention.

### 3 Classification Metrics Definitions

#### 3.1 Classification Metrics Terminology

The following terminology is used throughout this study to evaluate binary classification in the TACO framework:

- **True Positives (TP):** Correctly predicted Long COVID cases.
- **True Negatives (TN):** Correctly predicted non-Long COVID cases.
- **False Positives (FP):** Incorrectly predicted Long COVID cases (Type I error).
- **False Negatives (FN):** Missed Long COVID cases (Type II error).

#### 3.2 Primary Performance Metrics for TACO

**Precision (Positive Predictive Value):**

$$\text{Precision} = \frac{TP}{TP + FP} \tag{1}$$

Measures the proportion of predicted Long COVID cases that are actually positive. TACO achieved 69.2% precision, crucial for minimizing false positives in clinical deployment.

**ROC AUC (Area Under the Receiver Operating Characteristic Curve):**

$$\text{ROC AUC} = \int_0^1 \text{TPR}(FPR^{-1}(t))dt \tag{2}$$

Where:

- $\text{TPR} = \frac{TP}{TP+FN}$ (True Positive Rate)
- $\text{FPR} = \frac{FP}{FP+TN}$ (False Positive Rate)

The ROC AUC measures the model's ability to distinguish between classes at all classification thresholds. TACO achieved 0.663 ROC AUC, demonstrating superior discrimination capability [1].

**Additional Metrics.**

$$\text{Recall} = \frac{TP}{TP + FN} \tag{3}$$

Quantifies the proportion of actual Long COVID cases correctly identified (76.6% for TACO).

$$\text{F1-Score} = \frac{2 \times \text{Precision} \times \text{Recall}}{\text{Precision} + \text{Recall}} \tag{4}$$

Provides the harmonic mean of precision and recall (72.5% for TACO) [4].

### 4 Machine Learning Models: Equations and Rationale for Benchmarking

#### 4.1 TACO Framework: TabPFN with Causal Context

$$P(y_{test}|x_{test}^{causal}, D_{train}) = \text{Transformer}(x_{test}^{causal}, D_{train}, \Theta_{DCE})$$

Where

- $x_{test}^{causal}$ represents the 411 causal genes identified with DCE
- $D_{train} = \{(x_i, y_i)\}_{i=1}^{n}$ is the training set
- $\Theta_{DCE}$ encodes the causal context of the analysis of DCE.

#### 4.2 Benchmark Models (Using 500 Most Variable Genes)

**LINEAR MODELS Logistic Regression:**

$$P(Y = 1|X) = \frac{1}{1 + e^{-(\beta_0 + \sum_{i=1}^{500} \beta_i x_i)}} \tag{5}$$

**Ridge Classifier:**

$$\min_{\beta} \sum_{i=1}^{n} (y_i - \beta_0 - \sum_{j=1}^{500} \beta_j x_{ij})^2 + \lambda \sum_{j=1}^{500} \beta_j^2 \tag{6}$$

**Linear Discriminant Analysis (LDA):**

$$\delta_k(x) = x^T \Sigma^{-1} \mu_k - \frac{1}{2} \mu_k^T \Sigma^{-1} \mu_k + \log \pi_k \tag{7}$$

**TREE-BASED MODELS Random Forest:**

$$\hat{y} = \frac{1}{B} \sum_{b=1}^{B} T_b(x) \tag{8}$$

**Gradient Boosting:**

$$F_m(x) = F_{m-1}(x) + \gamma_m h_m(x) \tag{9}$$

**ADVANCED ENSEMBLE MODELS XGBoost:**

$$\mathcal{L} = \sum_{i=1}^{n} l(y_i, \hat{y}_i) + \sum_{k=1}^{K} \Omega(f_k) \tag{10}$$

**LightGBM:** Leaf-wise tree growth for efficiency with large gene sets.
**CatBoost:** Robust performance with minimal tuning.

**NEURAL NETWORK MODELS Multi-Layer Perceptron (MLP):**

$$h^{(l)} = \sigma(W^{(l)}h^{(l-1)} + b^{(l)}) \tag{11}$$

**Support Vector Classifier (SVC):**

$$\min_{w,b,\xi} \frac{1}{2}||w||^2 + C\sum_{i=1}^{n}\xi_i \tag{12}$$

#### 5 Experiment Configurations

TACO's configuration remained constant in all experiments:

- TabPFN with 411 DCE-identified genes.
- `device` = CPU (reproducibility).
- `N_ensemble_configurations` = 32.
- `no_preprocess_mode` = False.
- Gene weights proportional to $|\beta|$ from DCE.

##### 5.1 Experiment 1: Default Benchmark Hyperparameters

| Model | Key Parameters |
|---|---|
| Logistic Regression | `max_iter=1000`, `C=1.0` |
| Ridge Classifier | `alpha=1.0` |
| Linear Discriminant Analysis | `solver='svd'` |
| Random Forest | `n_estimators=100`, `max_depth=None` |
| Gradient Boosting | `n_estimators=100`, `learning_rate=0.1` |
| XGBoost | `n_estimators=100`, `eval_metric='logloss'` |
| LightGBM | `n_estimators=100`, `learning_rate=0.1` |
| CatBoost | `iterations=1000`, `learning_rate=0.03` |
| Multi-Layer Perceptron | `hidden_layer_sizes=(100,)`, `max_iter=1000` |
| Support Vector Classifier | `C=1.0`, `kernel='rbf'`, `probability=True` |

##### 5.2 Experiment 2: Tuned Benchmark Hyperparameters

| Model | Tuned Parameters |
|---|---|
| Logistic Regression | `C=0.5`, `penalty='l2'`, `solver='lbfgs'` |
| Ridge Classifier | `alpha=0.5` |
| LDA | `solver='lsqr'`, `shrinkage='auto'` |
| Random Forest | `n_estimators=500`, `max_depth=20`, `max_features='sqrt'` |
| Gradient Boosting | `n_estimators=500`, `learning_rate=0.05`, `max_depth=5` |

| | |
|---|---|
| XGBoost | `n_estimators=500`, `max_depth=6`, `learning_rate=0.05`, `subsample=0.8` |
| LightGBM | `n_estimators=500`, `max_depth=6`, `num_leaves=31`, `feature_fraction=0.8` |
| CatBoost | `iterations=1500`, `learning_rate=0.02`, `depth=6` |
| MLP | `hidden_layer_sizes=(200,)`, `alpha=0.0005`, `max_iter=2000` |
| SVC | `C=2.0`, `kernel='rbf'`, `gamma='scale'`, `probability=True` |

##### 5.3 Experiment 3: Strict CV Hygiene

| Model | Parameters (per-fold) |
|---|---|
| Logistic Regression | `max_iter=1000`, `C=1.0` |
| Ridge Classifier | `alpha=1.0` |
| Linear Discriminant Analysis | default settings |
| SVC | `C=1.0`, `kernel='rbf'`, `probability=True` |
| KNN | `n_neighbors=5` |
| MLP | `hidden_layer_sizes=(100,)`, `max_iter=1000` |
| Random Forest | `n_estimators=100`, `max_depth=None` |
| Gradient Boosting | `n_estimators=100`, `learning_rate=0.1`, `max_depth=None` |
| AdaBoost | `n_estimators=100`, `learning_rate=0.1` |
| Bagging | `n_estimators=100` |
| Extra Trees | `n_estimators=100`, `max_depth=None` |
| XGBoost | `n_estimators=100`, `learning_rate=0.1`, `max_depth=None` |
| LightGBM | `n_estimators=100`, `learning_rate=0.1`, `max_depth=None` |
| CatBoost | `iterations=100`, `learning_rate=0.1`, `depth=None` |
| Voting Classifier | Soft voting: LR, RF, XGB with above parameters |
| Stacking Classifier | Base: LR, RF, XGB; Final: Logistic Regression |

#### 6 Cross-Validation and Statistical Analysis

**Implementation:**

- Stratification maintains Long COVID prevalence across folds
- Random state=42 for reproducibility
- Each fold: ∼278 samples for testing, ∼1,114 for training

**Performance Improvements:**

- TACO Precision: 69.2% (±2.1%)
- Best Benchmark Precision: 66.3% (±3.8%)
- Improvement: +4.4% ($p < 0.01$, Wilcoxon signed-rank)

- TACO ROC-AUC: 0.663 (±0.023)
- Best Benchmark ROC-AUC: 0.602 (±0.041)
- Improvement: +10.1% ($p < 0.001$, Wilcoxon signed-rank)

**Effect Sizes:**

- Cohen's d for Precision: 0.92 (large effect)
- Cohen's d for ROC-AUC: 1.84 (very large effect)

### 7 Biological Validation of TACO's Causal Genes

Of the 411 genes identified by the DCE analysis of TACO, 97 genes (23. 6%) have established roles in the pathophysiology of Long COVID:
**Key Validated Genes:**

- **Viral Entry:** AR (androgen receptor), TMPRSS2, ACE2
- **Immune Response:** TP53, CDKN1A, RB1
- **Epigenetic Regulation:** CREBBP, EP300, HDAC1
- **Tissue Remodeling:** SMAD2, SMAD3 (TGF-$\beta$ pathway)

#### 7.1 Pathway Enrichment Analysis

Top enriched pathways among TACO's 411 causal genes:

- Viral infection response ($p < 0.001$)
- Inflammatory signaling ($p < 0.001$)
- Cell cycle regulation ($p < 0.01$)
- Epigenetic modification ($p < 0.01$)
- Tissue fibrosis pathways ($p < 0.05$)

### 8 Implementation and Reproducibility

**Core Dependencies:**

- Python 3.11
- TabPFN 0.1.0
- scikit-learn 1.3.0
- XGBoost 1.7.6, LightGBM 4.0.0, CatBoost 1.2
- pandas 2.0.0, numpy 1.24.0

**Computational Resources:**

- DCE Analysis: ~2 hours on 16-core CPU
- TabPFN Training: ~30 minutes per fold
- Total experiment time: ~4 hours

**Benchmark Models:**

- Training time: 5-60 seconds per model per fold
- Total for 16 models: ~2 hours

The complete TACO framework implementation is available at: https://github.com/AnonymousConferece/TACO
